# A Phase1 Results of a Non-Stabilized Spike-Encoding mRNA Vaccine in Adults

**DOI:** 10.1101/2022.05.12.22274989

**Authors:** Sivaporn Gatechompol, Wonngarm Kittanamongkolchai, Chutitorn Ketloy, Eakachai Prompetchara, Arunee Thitithanyanont, Anan Jongkaewwattana, Supranee Buranapraditkun, Mohamad-Gabriel Alameh, Sasiwimol Ubolyam, Jiratchaya Sophonphan, Tanakorn Apornpong, Stephen Kerr, Adeeba Kamarulzaman, Sarawut Siwamogsatham, Eugène Kroon, Thanyawee Puthanakit, Kanitha Patarakul, Tanapat Palaga, Wassana Wijagkanalan, Drew Weissman, Kiat Ruxrungtham, ChulaVAC-001 study team

**Author notes:** **Corresponding author** Kiat Ruxrungtham, MD, Center of Excellence in Vaccine Research and Development (ChulaVRC), Faculty of Medicine, Chulalongkorn University, 10^th^ Floor, Aor Por Ror Building, 1873 Rama IV Road, Pathumwan, Bangkok 10330, Thailand. Contributed equally as first author (Sivaporn Gatechompol, Wonngarm Kittanamongkolchai). Contributed equally as senior author (Drew Weissman, Kiat Ruxrungtham).

## Abstract

**Background:** Effective COVID-19 mRNA vaccines are mainly available in high-income countries. ChulaCov19, a prefusion non-stabilized Spike protein-encoding, nucleoside-modified mRNA, lipid nanoparticle encapsulated vaccine development, aims to enhance accessibility of mRNA vaccine and future pandemic preparedness for low- to middle-income countries.

**Methods:** Seventy-two eligible volunteers, 36 aged 18-55 (adults) followed by 36 aged 56-75 (elderly) enrolled in a dose escalation study of ChulaCov19 mRNA vaccine. Two doses of vaccine were given 21 days apart at 10, 25, or 50 µg/dose (12/group). Safety was the primary and immunogenicity the secondary outcome. Human convalescents’ (HCS) and Pfizer/BioNTech vaccinees’ sera provided comparison panels.

**Results:** All three doses of ChulaCov19 were well tolerated and elicited robust dose-dependent and age-dependent B- and T-cell responses. Transient mild/moderate injection site pain, fever, chills, fatigue, and headache were more common after the second dose. Four weeks after the second ChulaCov19: dose at 10, 25, and 50 µg dose, MicroVNT-50 Geometric mean titer (GMT) against wild-type was 848, 736 and 1,140 IU/mL, respectively, versus 267 IU/mL for HCS. All dose levels elicited 100% seroconversion, with GMT ratio 4-8-fold higher than for HCS (p<0.01), and high IFNγ spot-forming cells/million peripheral blood mononuclear cells. The 50 µg dose induced better cross-neutralization against Alpha, Beta, Gamma, and Delta variants than lower doses.

**Conclusions:** ChulaCov19 at 50 µg/dose is well tolerated and elicited higher neutralizing antibodies than HCS with strong T-cell responses. These antibodies cross neutralized four variants of concern and ChulaCov19 has therefore proceeded to phase 2 and 3 clinical trials.

**Trial registration number:** ClinicalTrials.gov Identifier NCT04566276

**Graphical Abstract:** 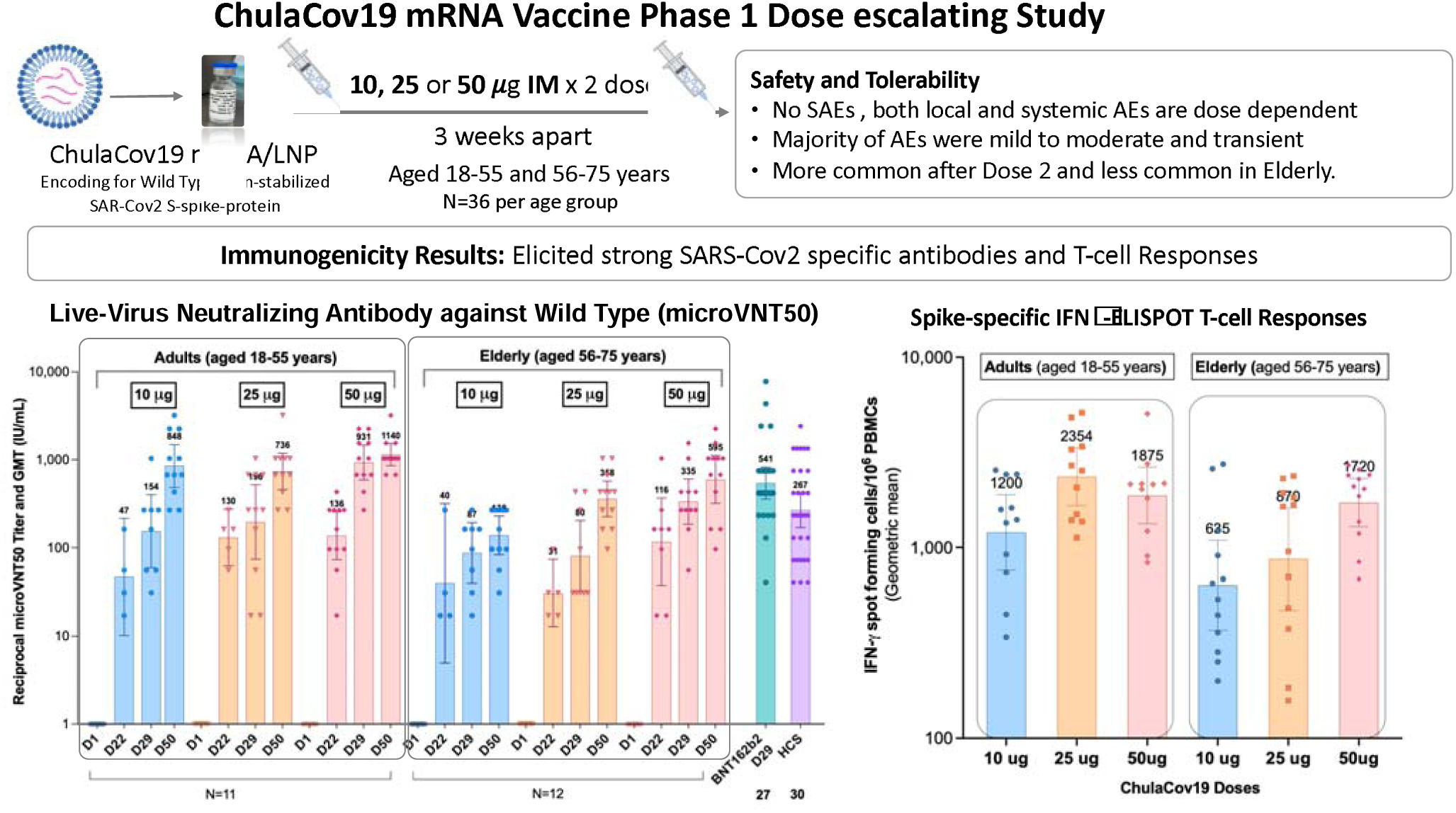

## INTRODUCTION

More than 10 billion doses of COVID-19 vaccines have been administered worldwide, however only 10 % of those living in low-income countries received at least one dose (8 February 2022)^1^. Among approved vaccines, mRNA vaccines have shown the highest efficacy^2^. Building capacity to develop and manufacture mRNA vaccines in low- to middle-income countries (LMIC) is important for the current and future pandemics. The ChulaCov19 mRNA vaccine (ChulaCov19) is a lipid nanoparticle (LNP) encapsulated nucleoside-modified mRNA, encoding a non-stabilized, full-length SARS-Cov2 spike protein. Two major differences between ChulaCov19 with the Pfizer/BioNTech and Moderna vaccines are that ChulaCov19 is not prefusion stabilized and is encapsulated in a different LNP formulation. ChulaCov19 was conceptualized in Thailand and initially manufactured in North America for early clinical development, in parallel with large-scale production capacity development in Thailand. In rodent and macaques, ChulaCov19 elicited strong B and T cells responses. The induced high neutralizing antibody levels against wild-type virus were able to cross neutralize four variants of concern (VOC). Here, we report a phase 1 open-label, dose-escalation study of ChulaCov19.

## Methods

### Trial Design

This phase 1, open-label, dose escalation study to evaluate safety and immunogenicity of ChulaCov19 vaccine enrolled healthy participants aged 18-55 (adults, n=36) followed by a cohort aged 56-75 (elderly, n=36). The inclusion and exclusion criteria are shown in the **Supplemental Table S1**. Both age cohorts received ChulaCov19 at 10, 25, or 50-µg per dose, 12 participants per dose group for each age cohort, in a sentinel, dose-escalating manner.

The trial and the Investigational New Drug application were approved by the ethics committee of the Faculty of Medicine, Chulalongkorn University, Bangkok, and Thailand’s Food and Drug Administration, respectively. All participants provided written informed consent. The trial was conducted at the Chula Clinical Research Center and King Chulalongkorn Memorial Hospital, Bangkok.

### Trial vaccine

ChulaCov19 mRNA was manufactured at Trilink Biotechnologies (San Diego, California), and Integrity Bio Inc. (Camarillo, California). The encapsulating LNP formulation consists of four lipid components with a proprietary ionizable ionized lipid developed by Genevant Sciences Corporation (Vancouver, British Columbia). ChulaCov19 is stable at -75±10°C, -20±5°C, and, importantly, 5±3°C for up to 6 months. ChulaCov19 was stored as a sterile suspension of 0.2 mg/ml at -75±10°C and diluted with normal saline according to the assigned dose, to be given at 0.5 ml intramuscularly (IM).

### Trial procedures

Four adult sentinel participants were enrolled to receive ChulaCov19 at 10-µg (IM). Once no halting criteria **(Table S2)** were reported by Day 3, the remaining eight 10-µg participants were enrolled. The same approach was followed for the 25- and 50-μg doses. The vaccine was administered in the deltoid muscle on Day 1 and Day 22±3, followed by 2 hours safety monitoring on-site. Enrollment of the elderly participants was commenced after DSMB review of the data when the last adult participant of the 10-µg group had reached Day 29 (one week after second vaccination) on study. A diary was provided to participants to record solicited- and unsolicited adverse reactions, and concomitant medications for seven days. Safety laboratory tests were performed at baseline, and days 8, 22, 29 and 50.

### Assessment of safety and tolerability

Safety endpoints included solicited and unsolicited local adverse events (AE), systemic AE, use of antipyretics/analgesics in the seven days after vaccination, and serious adverse events (SAE) up to Day 50 (four weeks after second vaccination).

### Assessment of immunogenicity

The binding to the SARS-CoV-2 S1 receptor-binding domain (RBD) was measured by enzyme-linked immunosorbent assay (ELISA). Neutralizing antibody titers against wild-type and VOC were assessed by live-virus microneutralization assay (MicroVNT-50) and pseudovirus neutralization assay (PsVNT-50). SARS-Cov2 RBD-ACE2 blocking antibody was measured by surrogate viral neutralization test. Cellular immunity was measured by IFNγ-ELISpot assay (ELISpot), and by intracellular cytokine staining (ICS) assay **(Table S3)**. Tests were performed on specimens collected at Days 1, 8, 22, 29 and 50; however, PsVNT-50 was performed only at Days 29 and 50. Exploratory comparator serum panels included 30 human convalescents’ serum (HCS) from adults with COVID-19; serum samples were collected at four weeks after confirmed diagnosis, and 27 Pfizer/BioNTech mRNA vaccinees; serum samples were collected at Day 29 **(Table S4 and S5)**.

### Statistical Analysis

Sample size for Phase I was based on practical and medical considerations, not power for statistical hypothesis testing or precision of parameter estimation. Results of safety analyses are presented as counts, percentages and group ages as mean (SD). Summary descriptive statistics relevant for study endpoints were provided for each cohort and vaccine dose group at the study time points indicated in the protocol.

Immunogenicity was analyzed based on the per protocol (PP) analysis set. Geometric mean titers (GMT) and their 95%CI were calculated for MicroVNT-50, PsVNT-50, anti RBD-IgG, anti S-Trimer, sVNT, IFNγ-ELISpot and Th1/Th2 spike-specific-CD4+ and CD8+ T-cells. Formal comparisons of MicroVNT-50, PsVNT-50, anti RBD-IgG and sVNT against samples of 30 HCS from adults with COVID-19 and 27 Pfizer/BioNTech mRNA vaccinees, were made using geometric mean ratios (GMR 95%CI). Statistical analysis was conducted with Stata 15 (Statacorp LLC, College Station, TX, USA).

## Results

Between 28 May 2021 and 2 July 2021, 132 individuals were screened of whom 36 were enrolled in each age cohort (12 per dose group in each age cohort) (**Figure S1 and S2**). All but one participant completed both vaccinations. The participant declining second vaccination was from the adult 25-μg group and experienced moderate myalgia after the first vaccination, which resolved within three days and which was accompanied by moderate CPK elevation but normal cardiac enzymes, possibly related to strenuous physical labor. This participant was excluded from the per protocol immunogenicity analysis as were two others: one participant from the adult 10-μg group who was diagnosed with asymptomatic COVID-19 on the sixth day after the second vaccination and one participant from the adult 50-μg group who had confirmed positive baseline anti-RBD binding antibody test but was negative for SARS-CoV-2 molecular testing, indicating asymptomatic COVID-19 prior to enrolment. All participants contributed to the safety analysis.

### Characteristics of the study participants

Characteristics of the study participants are listed in **Table 1**. Mean age (SD) in the adult and elderly cohorts was 35.8(9.1) and 65.1(5.4) years, respectively; body mass index was 23(2.8) and 23.3(3.3); and proportion of females was 67% and 53%.

**Table 1:**
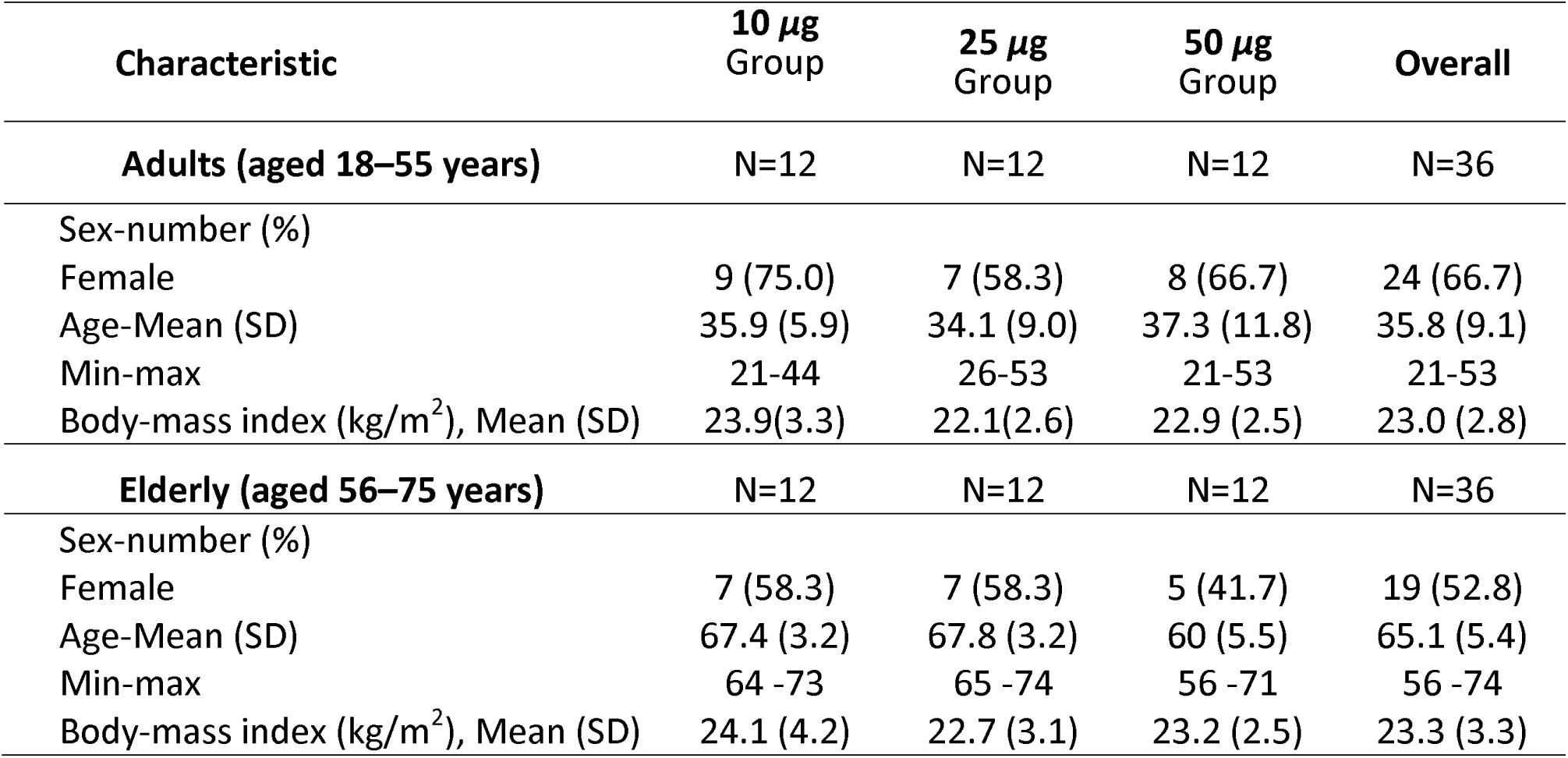
Demographic Characteristics of the participants by dose age groups

### Safety and Tolerability

#### *Local reactions* (Figure 1)

The most common local reaction was injection site pain. Its incidence was dose-dependent, more common in adults than elderly, and after Dose 2. All local reactions were mild in the elderly; some moderate events occurred in adults. All participants recovered after 2.79±1.7 and 1.91±0.9 days on average in adults and elderly after Dose 2, respectively. A single event of severe erythema in the adult 10-μg group after Dose 2 resolved within 6 days.

**Figure 1.**
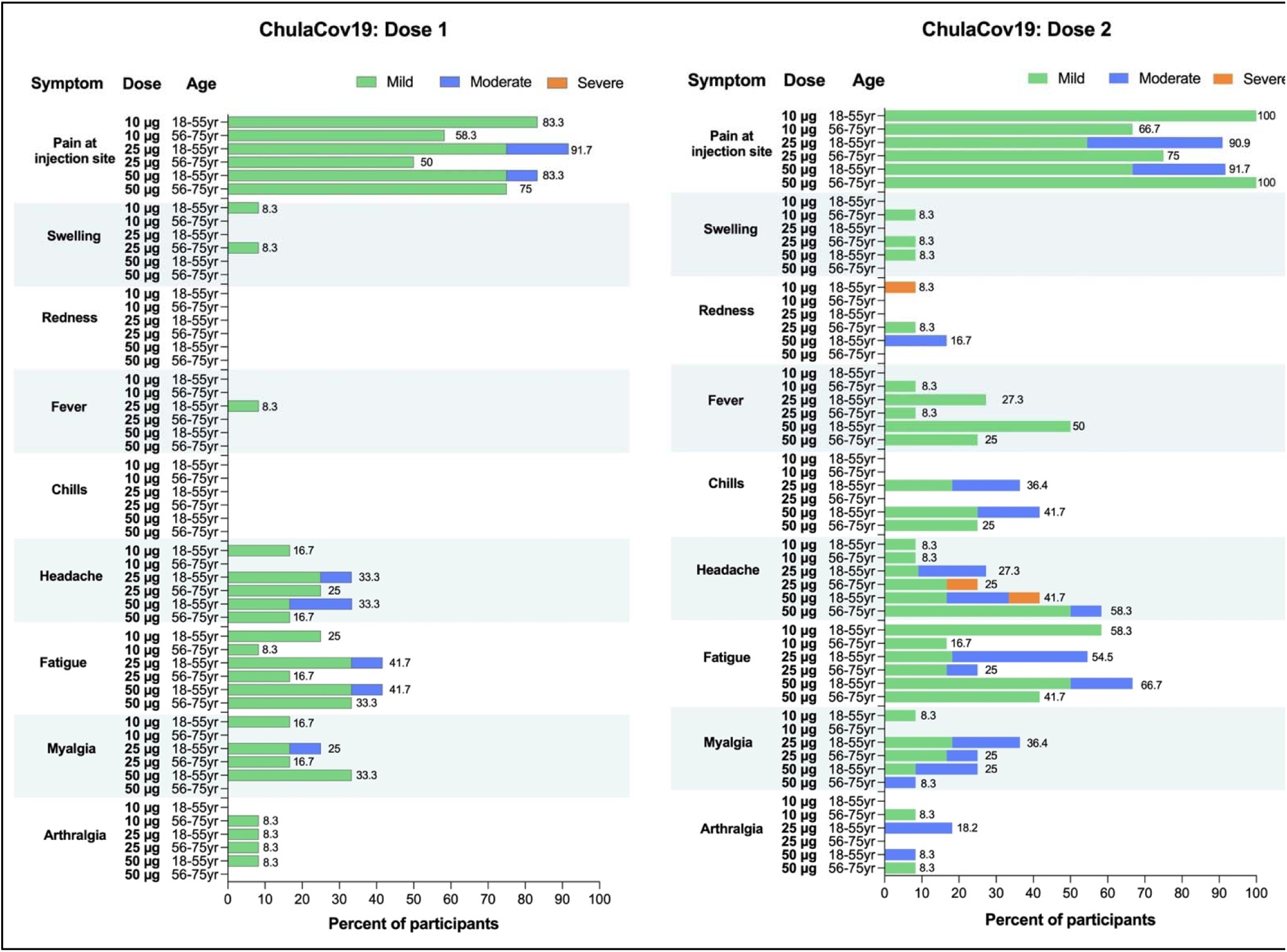
Local and Systemic Adverse Events, by dose group and age cohorts. The 18-55- and 56-75-years old cohort is also referred to as adults and elderly.

#### *Systemic reactions* (Figure 1)

The three most common systemic reactions were fever, headache, and fatigue. Systemic reactions were more common at the 25-μg and 50-μg doses, after Dose 2, and in adults. Most reactions were mild to moderate and transient with mean duration 1.97±1.2 days in adults and 1.39±0.5 days in elderly after Dose 2.

### Immunogenicity Results

#### SARS-CoV-2 Receptor Binding Domain (RBD) Antibody Responses (Figure 2a)

In the adult cohort, 4 weeks after second vaccination (Day50) the GMT of anti-RBD antibody was significantly higher than for HCS 4 weeks after COVID-19 diagnosis with GMR of 3.78, 3.23, and 7.74, p≤0.01, respectively. In the elderly cohort, anti-RBD GMT were also dose-dependent but lower than observed in the adult cohort **(Table S6)**.

**Figure 2.**
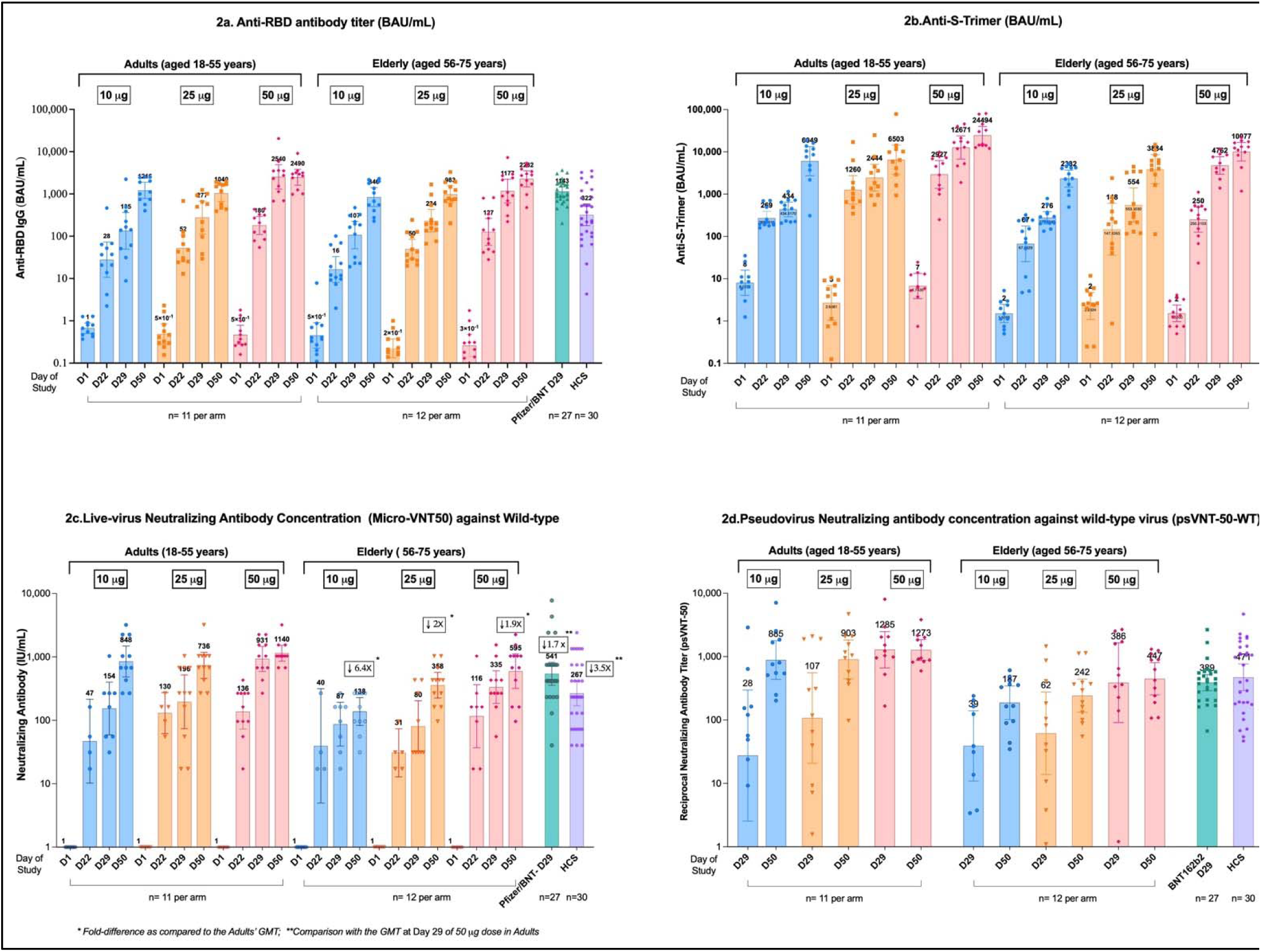
SARS-CoV-2 Antibody-Binding and Neutralization Responses.

#### SARS-CoV-2 S-Spike Trimer Antibody Responses (Figure 2b)

In the adult cohort at Day 50, ChulaCov19 at 50-ug elicited highest anti-Spike-GMT at 24493.9 BAU/mL (p<0.01). In the elderly cohort, the anti-Spike GMT was lower than for the adults at all three doses while anti Spike-GMT for the 50-µg dose was significantly higher than for the 10-µg dose, p 0.001.

#### RBD-ACE2 Binding Inhibition Antibody Responses (sVNT)

GM of % RBD-ACE2 binding inhibition showed a dose-dependent pattern in both age-cohorts at Day 29 but not at Day 50. All doses at Day 50 in both age-cohorts elicited GM>90% inhibition. % Inhibition of the HCS and Pfizer/BNT vaccinee’s (Day 29) panels was 76% and 93%, respectively **(Figure S3)**.

#### *SARS-CoV-2-*specific Live-virus Micro-Neutralization Tests (MicroVNT-50)

Seroconversion rate on MicroVNT-50 in all dose groups reached 100% by Day50. GMT of the responses against WT virus were both dose- and age-dependent, and markedly increased from Day 29 to Day 50 for the 10, 25 and 50 µg doses **(Figure 2c)**. In the adult cohort at Day50, ChulaCov19 at 10, 25 and 50-µg doses induced significantly higher MicroVNT-50 GMT than HCS at a ratio of 2.98, 2.59 and 4.01, respectively (p<0.01), whereas in the elderly cohort the values were: 0.48, 1.26and 2.09-folds, respectively. **(Table S6)**. ChulaCov19 at 50-µg dose induced significantly higher MicroVNT-50 GMT against wild-type and 3 variants as compared to the 10-µg dose in both age cohorts while in the elderly cohort elicited MicroVNT-50 GMT were lower for all doses when compared to the adult cohort. **(Figure 3a and Table S7)**.

**Figure 3.**
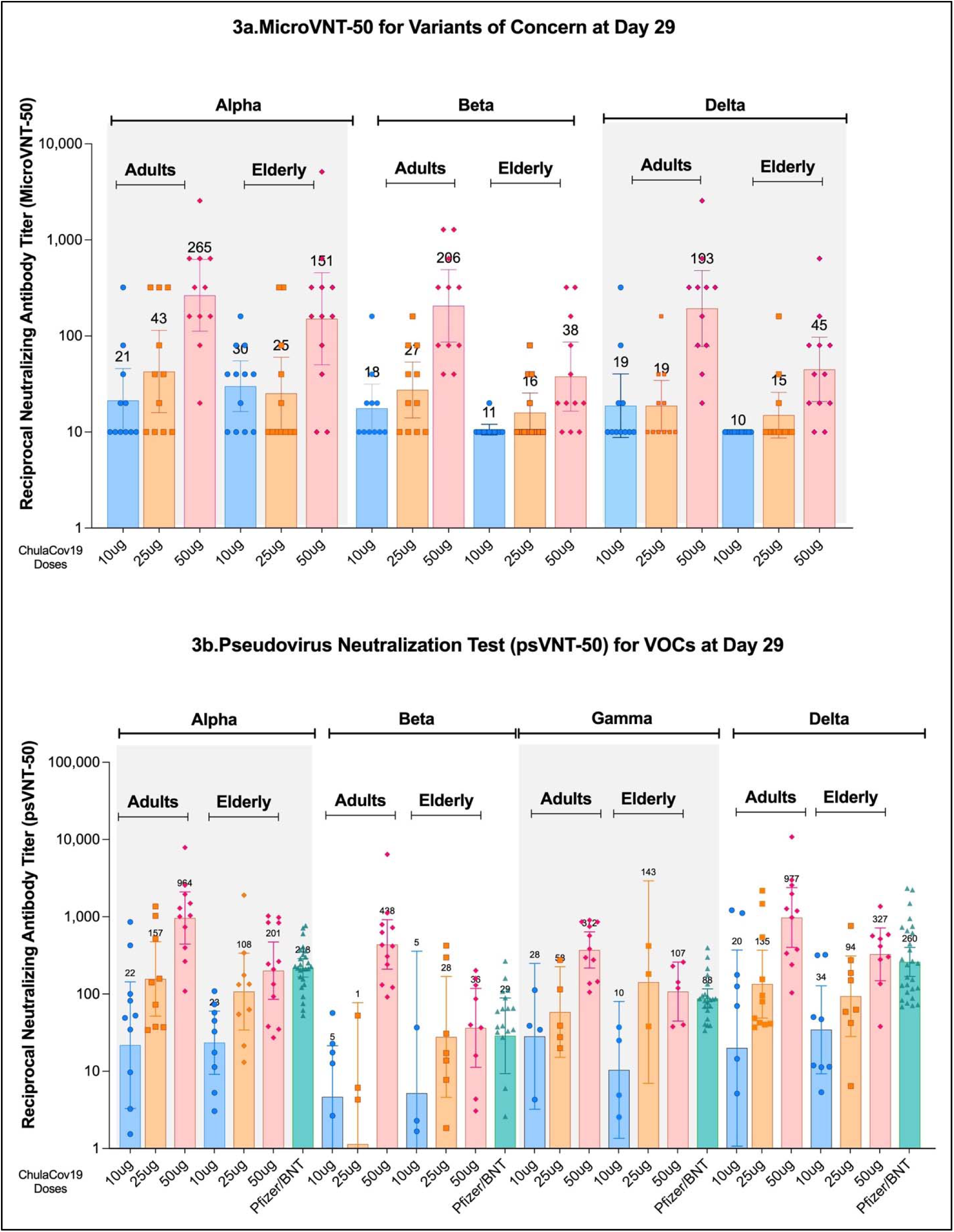
SARS-CoV-2 Neutralization Responses against variants of concern at Day 29 (4 weeks after Dose2)

#### Pseudovirus Neutralization Tests

In the adult cohort, PsVNT-50 GMT (95% CI) of ChulaCov19 against WT at Day50 at 10, 25, 50-µg and of HCS panel was 885.3, 902.9, 1,273.1, and 471.2, respectively. (**Figure 2d**). The GMT-ratio of 50 µg dose/HCS was 2.7 (p=0.01). At Day 29, ChulaCov19 at 50-µg dose induced significantly higher GMT than the Pfizer/BNT vaccinated panel against Alpha, Beta and Gamma variants at a ratio of 4.47, 11.94 and 4.23 (p=0.01), respectively (**Figures 3b and Table S8**). A new variant Omicron PsVNT-50 test results showed two-doses of ChulaCov19 at 50-µg has a significant decline of GMT against Omicron variant as compared to WT (**Figure S4**).

#### SARS-Cov2 Spike-specific T-Cell Responses

At Day 29, all participants in the adult and elderly cohorts at 10-, 25-, and 50-µg showed strong Spike-specific T-cell responses measured by IFNγ-ELISpot tests. The responses were lower in elderly at 10- and 25-ug than in adults (**Figure 4a and Table S9**). In adults, both Spike-specific IFNγ^+^CD4^+^ and IL2^+^-CD4^+^ T-cell percentages were significantly higher in the 25- and 50-µg dose recipients when compared to the 10-µg dose. **(Table S10)**. ChulaCov19 elicited Spike-specific Th1-dominated responses (**Figure 4b**).

**Figure 4:**
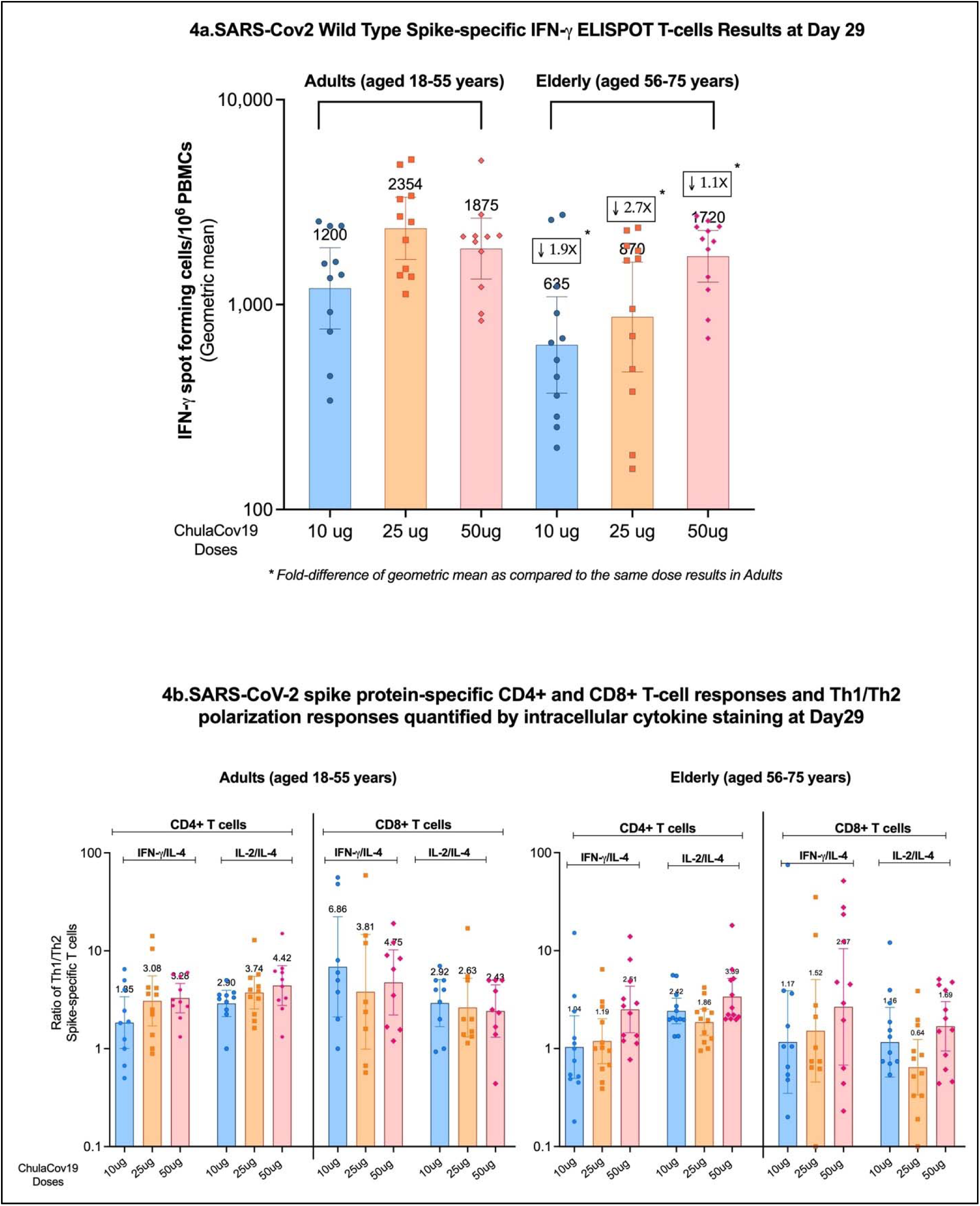
SARS-Cov2 Wild-type Spike-specific T-cells responses

## Discussion

To enhance access to effective COVID-19 mRNA vaccines in LMIC and to be prepared for a next pandemic, complete mRNA-LNP vaccine development and manufacturing value chains need to be established in LMICs. Unlike currently approved mRNA vaccines, such as Pfizer/BNT and Moderna^3^, ChulaCov19 is a wild-type non-stabilized Spike-protein encoded mRNA vaccine encapsulated with a different LNP and demonstrated thermostability at 2-8° C for up to 6 months. In this phase 1 trial, three doses of ChulaCov19 vaccine were well tolerated with no SAE observed in either age-group. Injection site pain is the most common AE while fever, chills, headache, and myalgia were reported to be dose-dependent and more frequent after the second dose. AEs were both less frequent and milder among the elderly participants. Moderate to severe events were rare overall and all AEs resolved on average 2.5 days.

The results indicated that this LNP-encapsulated non-di-Proline-stabilized Spike-protein mRNA vaccine is strongly immunogenic. ChulaCov19 at 50-µg dose induced high SARS-CoV2-binding and neutralizing antibodies one week after dose 2 (p<0.01) with a microVNT-50 GMT 6-fold higher than for HCS. At 4 weeks after dose 2, there was a further rise of all tested antibodies and all 3 doses elicited microVNT-50 GMT higher than HCS with a GMT ratio of 4-8-fold (p<0.01). The 50-µg dose induced higher micro-VNT-50 and psVNT-50 GMT against tested variants than the lower doses (p<0.01). In the adult cohort, at one week after Dose 2, the ratios of ChulaCov19 50-µg dose elicited psVNT-50 GMT against Alpha, Beta, and Gamma variants that were 4.5, 11.9 and 4.2, respectively higher compared to Pfizer/BNT vaccine(p<0.01). In term of Omicron variant, ChulaCov19 vaccine, as reported in approved Covid-19 mRNA vaccine, 2-doses vaccination may not effective against Omicorn variant, a Third dose is required^4^. Developing pan-SARS-Cov2 or pan-Coronavirus vaccine against future pandemic by is warranted^5^.

All doses of ChulaCov19 generated strong T cell responses and the higher doses (25 and 50-µg) elicited higher % of Spike-specific IL2+CD4+T-cells and % of IFNγ+CD4+T-cells than the 10-µg dose (p<0.01 and p<0.05, respectively). All doses ChulaCov19 elicited predominantly SARS-CoV2-specific Th1-type responses. Based on these results, the DSMB has recommended that ChulaCov19 be further advanced to a phase 2 randomized-controlled trial with the 50-µg dose.

This vaccine did not contain prefusion stabilization of Spike with 2 prolines (K986P/V987P) that many approved COVID-19 vaccine platforms use^6^. The Phase I data presented here show ChulaCov19 elicited robust humoral and T cell responses, which are often greater than the BioNTech/Pfizer vaccine that contains the di-proline modification. There are other differences in the BioNTech/Pfizer modified mRNA vaccine that could account for the greater potency of ChulaCov19, including different UTRs, coding sequence optimization, poly(A) tail, and the LNP formulation used. Our data does support that the Spike does not need to be prefusion stabilized to induce a potent and protective response, unlike RSV^7^.

Recent studies suggest that a GMT-ratio of neutralizing or binding antibody in vaccinees to the level in convalescent patients of >1 is associated with >70% vaccine efficacy rate (VE).^2^ In addition, higher binding and neutralizing antibody levels at four weeks after the second dose were found to correlate with symptomatic infection risk reduction in the AZD1222 trial^8^ and COVE study^9^. ChulaCov19 induced strong binding and neutralizing antibody responses four weeks after the second dose and the ratio of neutralizing antibody GMT in ChulaCov19 vaccinees vs convalescent sera of four weeks after diagnosis of approximately four to eight folds suggests that this vaccine candidate has a potentially significant vaccine efficacy. A consensus on correlates of protection may be achieved soon^10^ and recently the International Coalition of Medicines Regulatory Authorities (ICMRA) has accepted well-designed immunobridging studies for authorizing COVID-19 vaccines^11^.

Accessibility to effective COVID-19 vaccines, particularly mRNA vaccines, remains very limited in many low-middle income countries (LMICs)^12^. The main goal of ChulaCov19 development is to be part of the solution for LMICs. There are several challenges to overcome: the speed of clinical development to reach emergency use authorization (EUA), the speed of establishing large scale manufacturing capacity, negotiation of expanding LNP-licensing territory, and the cost and long lead-time raw materials. Currently Bionet Asia, Thailand has already established manufacturing capacity for both mRNA production and encapsulation, and a first clinical lot has been released for further clinical development and EUA approval.

The limitations of this study include: the sample size is small due to the phase 1 uncontrolled dose-finding design. The exploratory comparative immunogenicity analyses with convalescent sera or Pfizer/BNT vaccinees’ sera are not direct head-to-head comparisons and can contain bias. Convalescent sera were collected during the rise of the Delta variant outbreak, and possibly antibody responses are stronger against Delta than WT. To minimize bias, the convalescent and Pfizer/BNT vaccinees’ serum samples were tested at the same laboratories together with the ChulaCov19 vaccinated samples. In addition, a RCT phase 2 study has commenced and a larger scale immune-bridging, non-inferiority phase 3 study is planned.

In summary, ChulaCov19 mRNA vaccine is well tolerated and elicited strong SARS-CoV2 specific B- and T-cell immunogenicity and is currently under Phase 2 and later clinical development.

## Data Availability

All data produced in the present work are contained in the manuscript

## Acknowledgement

This study was funded by National Vaccine Institute (NVI), grant no. 2563. 1/ 11 and 2564.1/4; Chulalongkorn University Second Century Fund (C2F); the Ratchadapisek Sompoch Endowment Fund (2021), Chulalongkorn University (764002-HE04); and Public Donation through Covid-19 vaccine development fund of the Faculty of Medicine, Chulalongkorn University and the Thai Red Cross Society, Thailand. The authors would like to thank all the study participants without whose support the study would not have been possible.

