## Supplementary appendix for "A Phase1 Results of a Non-Stabilized Spike-Encoding mRNA Vaccine in Adults"

This appendix has been provided by the authors to give readers additional information about their work.

### Table of contents:

| Supplementary Appendix | Page |
|---|---|
| 1. **ChulaVAC-001** Study Team and Affiliations | 3 |
| 2. **Table S1:** The study inclusion and exclusion criteria | 4-6 |
| 3. **Table S2:** The study halting criteria | 7 |
| 4. **Table S3:** The Immunogenicity laboratory methods | 8-10 |
| 5. **Table S4:** Demographic characteristics of human convalescent serum samples | 11 |
| 6. **Table S5:** Demographic characteristics of Pfizer/BioNTech mRNA vaccinees | 11 |
| 7. **Figure S1:** Enrollment and Distribution Adult Cohort | 12 |
| 8. **Figure S2:** Enrollment and Distribution Elderly Cohort | 13 |
| 9. **Table S6**: Comparison of Anti RBD-IgG Antibody (BAU/mL) at Day 29 and 50 Results of ChulaCov19 Vaccine 25 and 50 μg Dose with 10 μg Dose as Reference | 14 |
| 10. **Figure S3:** Surrogate Viral Neutralizing Antibody assay (sVNT) | 15 |
| 11. **Table S6**: Comparison of SARS-COV-2-specific Serum Neutralizing Antibody as Measured by Live-Virus Micro-VNT50- Against Wild-type, Alpha, Beta, and Delta at Day 29 and 50 of ChulaCov19 Vaccine Doses with 10 μg as a Reference. | 16 |
| 12. **Table S7**: Comparison of MicroVNT-50 Results of Adult Aged 18-55 against SARS-Cov2 Wild-Type virus and Variants of Concerns between ChulaCov19 vaccine and human Convalescent Serum Panel as a Reference (GMTR= GMT ratio) | 17 |
| 13. **Table S8:** Comparison of Pseudovirus Neutralizing Antibody (psVNT-50) Results of ChulaCov19 Vaccine with Pfizer /BNT Vaccine as a Reference | 18 |
| 14. **Figure S4** Pseudovirus neutralizing antibody results against Omicron variant of Adults and Elderly vaccinated with ChulaCov19 vaccine at 50 μg dose | 19 |
| 15. **Table S9:** Comparison of IFN-γ-ELISPOT T Cell responses at Day 29 of ChulaCov19 Vaccine Doses with 10 μg as a Reference. | 20 |
| 16. **Table S10**: Comparison of percent of SARS-Cov2-Spike-specific-CD4+ T-cell responses and Th1/Th2 ratio measured at day 29 by intracellular cytokine-staining assays | 21 |

### ChulaVAC-001 Study Team and Affiliations

Suwimon Manopwisedjaroen, M.Sc.[1], Thanida Laopanupong, M.Sc.[1], Supanuch Ekronarongchai, M.Sc.[1], Chanya Srisaowakarn, M.Sc.1, Yuparat Jantraphakorn, B.S.[2], Kanjana Srisutthisamphan, M.Sc.[2], Reena Rajasuriar, Ph.D[3], Supalak Phonphithak, Ph.D[4], Jutatip Sunjirat, M.Sc.[4], Apicha Mahanontharit, B.Sc.[4], Bunruan Sopa, B.Sc.[4], Nuchthida Phongam, B.Sc.[4], Umaporn Chobkarching, M.Sc.[4], Siriwan Thongthip, M.Sc.[5], Konsiri Soisoongnern, M.Sc.[5], Chomnid Shanyip, M.Sc.[5], Thanakan Rachpradit , B.Sc.[5], Peepattra Wantanasiri, Ph.D.[5], Khunthalee Benjapornpong, BNS.[6], Kamonkarn Tungnaree, MA.[6], Ponsuk Visudhipan Grandin, M.Sc.[7]

### Affiliations

1. Department of Microbiology, Faculty of Science, Mahidol University, Bangkok, Thailand
2. Virology and Cell Technology Research Team, National Center for Genetic Engineering and Biotechnology (BIOTEC), National Science and Technology Development Agency (NSTDA), Pathumthani, Thailand.
3. Department of Pharmacy, Faculty of Medicine, University of Malaya
4. HIV-NAT, Thai Red Cross AIDS Research Centre, Bangkok, Thailand
5. Clinical Research Center (Chula CRC), Faculty of Medicine, Chulalongkorn University, Bangkok, Thailand
6. SEARCH Projects, Institute of HIV Research and Innovation (IHRI), Bangkok, Thailand
7. Armed Forces Research Institute of Medical Sciences, Bangkok, Thailand

### Table S1: The study inclusion and exclusion criteria

**Inclusion Criteria**

Participants who meet all the following criteria at Screening are eligible to participate in the study:

All participants:

1. Participants must be able to communicate effectively with study personnel and considered reliable, willing, and cooperative in terms of compliance with the protocol requirements.

2. Participants must sign the written informed consent form prior to undertaking any protocol related procedures.

3. Participants must have a body mass index (BMI) at Screening, calculated as the body mass divided (in kilograms [kg]) by the square of the body height (in metres [m]) of 18.0-30.0 kg/m2, inclusive.

4. Participants must have haematology, clinical chemistry, coagulation (for all participants in Phase 1, and, only if applicable, for participants in Phase 2), and urinalysis test results that are not deviating from the normal reference range by age and gender to a clinically relevant extent at Screening.

5. Males must be surgically sterile (>30 days since vasectomy with no viable sperm), practice true abstinence or, if engaged in sexual relations with a female of child-bearing potential, the participants and their partner must use an acceptable, highly effective, double-barrier contraceptive method* from Screening and for a period of at least 60 days after the last dose of investigational vaccine.

6. Women of child-bearing potential must practice true abstinence or, if engaged in sexual relations with a male, they must agree to use highly effective (failure rate of < 1% per year when used consistently and correctly), double-barrier contraceptive measures* throughout the study and intend to continue use of contraception for at least 60 days following the last vaccination.

* The PI is to assess the adequacy of methods of contraception on a case-by-case basis (please consult Appendix 1 for further details). These criteria do not apply if the participants are in a same-sex relationship.

7. Women of child-bearing potential must have a negative serum pregnancy test (beta human chorionic gonadotropin [β-HCG]) at Screening and a negative urine-based test within 24 hours prior to each investigational vaccine administration.

8. Women of non-child-bearing potential must:

a. be classified as being postmenopausal (defined as having a history of amenorrhea of at least one year), or

b. where history of amenorrhea is less than one year, female participants must have a follicle stimulating hormone (FSH) level > 40 milli-international units per millilitre (mIU/mL), or

c. have a documented status of being surgically sterile (hysterectomy, bilateral oophorectomy, or tubal ligation/salpingectomy).

9. Participants must be in general good health based on medical history and physical examination, as determined by the PI (please consult Appendix 1 for further details), at Screening.

10. Body temperature must be less than 37.8 C, at Screening.

11. Pulse must be no greater than 100 beats per minute, at Screening.

12. Systolic blood pressure (SBP) must be between 85 to 150 millimetres of mercury (mm Hg), inclusive, at Screening.

13. Participants must agree to refrain from donating blood, plasma, ovules, sperm, or organs during the whole study.

<u>Adult Participants (Group 1 of Phase 1) only</u>

14. Must be a male or female aged 18-55 years (inclusive) at the time of enrolment.

<u>Elderly Participants (Group 2 of Phase 1) only</u>

15. Must be a male or female aged 56-75 years (inclusive) at the time of enrolment.

**Exclusion Criteria**

The presence of any of the following criteria will constitute cause for the exclusion of the participant:

1. Presence of clinically significant medical history, unstable chronic or acute disease, or physical, or laboratory findings that, in the opinion of the PI may potentially increase the expected risk of exposure to the investigational vaccine, compromise the safety of the participant, or interfere with any aspect of study conduct or interpretation of results. This will include asthma and any thrombocytopenia or bleeding disorder contraindicating IM vaccination (please consult Appendix 1 for further details).

2. Presence of self-reported or medically documented significant medical or psychiatric condition(s).

3. Presence of an acute illness, as determined by the participating site PI or appropriate sub-PI (please consult Appendix 1 for further details), with or without fever ( temperature ≥ 38.0 C) within 72 hours prior to each vaccination.

4. Presence of birthmarks, tattoos, wound, or other skin conditions over the deltoid region of both arms that, in the PI's opinion, could reasonably obscure and interfere with evaluation of local ISRs.

5. Inadequate venous access to allow collection of blood samples.

6. Breastfeeding or planning to breastfeed from the time of the first vaccination through 60 days after the last vaccination, or pregnant as confirmed by a positive serum β-HCG pregnancy test at Screening or positive urine pregnancy test at subsequent clinic visits at timepoints as delineated in the schedule of assessments.

7. Received any prophylactic or therapeutic vaccine, or licensed or unlicensed vaccine, drug, biologic, device, blood product, or medication, within 4 weeks of first vaccination or 5 half-lives (whichever is longer)

8. Participant has previously participated in an investigational study involving LNPs (a component of the investigational vaccine assessed in this trial).

9. History of severe allergy (requiring hospital care), severe reaction to any drug or prior vaccination, or any known or suspected allergies or sensitivities to any component of the investigational vaccine or placebo.

10. History of ever had an anaphylaxis reaction to food, medication or vaccination.

11. Participant is immunosuppressed as caused by disease (such as HIV).

12. Chronic use (more than 14 continuous days) of or anticipated need to use, within the next 6 months, of any medications that may be associated with impaired immune responsiveness or with immunosuppression.

13. History of hepatitis B or hepatitis C infection.

14. Receipt of immunoglobulins or blood products within 3 months of first vaccination.

15. Requirement for antipyretic or analgesic medication on a daily or every other day basis from enrolment through 72 hours after vaccination.

16. Current use of any prescription or over-the-counter medications within 7 days prior to vaccination, unless approved by the PI.

17. History of alcohol or drug abuse that in the opinion of the PI could affect the participant's safety or compliance with study.

18. Participant unwilling to abstain from blood donation during the course of the study, and/or participation in any research study involving blood sampling (more than 450 mL /unit of blood), or blood donation to any blood bank during the 2 months prior to the Screening visit.

19. Close contact with anyone known to have SARS-CoV-2 infection within 30 days prior to vaccine administration.

20. Positive on SAR-CoV-2 antibody IgG/IgM and anti spike IgG at screening

21. History of COVID-19 diagnosis (the criteria for COVID-19 diagnosis will follow the local guidelines).

22. On current treatment with investigational agents for prophylaxis of COVID-19.

23. Planning to travel outside Thailand from enrolment through 28 days after the second vaccination.

24. Residing in a nursing home or other skilled nursing facility or having a requirement for skilled nursing care.

25. Is a participant at high risk of SARS-CoV2 exposure in the opinion of the PI (e.g., healthcare workers, active health care workers with direct patient contact, emergency response personnel).

<u>Elderly Participants (Group 2 of Phase 1) only</u>

26. Chronically smoking (defined as ≥10 Pack years [packs/day years smoked]) within the 12 months prior to enrolment.

27. Presence of co-morbidities that can be associated with an increased risk of severe COVID-19; Cancer, Chronic kidney diseases, COPD, cardiovascular disease, solid organ transplantation, DM type 2, HT, cerebrovascular disease, Obesity (BMI> 30 kg/$m^2$)

### Table S2: The study halting criteria

Halting Criteria for Sentinel Participants

- Any participant experiences any event of ulceration, abscess, or necrosis at the injection site.
- Any participant experiences any event of laryngospasm, bronchospasm, or anaphylaxis within 24 hours after study vaccine injection.
- Any participant experiences 7eutralizin urticaria (defined as occurring at 3 or more body parts) within 72 hours after administration of vaccine.
- Any participant experiences an SAE (except for accident or trauma) after administration of the vaccine that is considered related to the vaccine.
- Any 2 participants in the same cohort experience the same Grade 3 Solicited Local AE or Systemic AE, (excluding measured grades of erythema and oedema/induration alone) that lasted at least 48 hours within 7 days after administration of the vaccine.
- Any 2 participants experience the same Grade 3 AE (unsolicited and/or clinical laboratory abnormality), in the same PT based on the MedDRA coding, that lasted at least 48 hours after administration of the vaccine and is considered related to the vaccine. Clinical laboratory abnormalities are not subject to the time window.

Halting Criteria for All Participants Except Sentinels

- Any participant experiences an SAE within 28 days after vaccine injection, that is considered related to the vaccine.
- Any participant experiences any event of laryngospasm, bronchospasm, or anaphylaxis within 24 hours after study vaccine injection, that is considered related to the vaccine.
- Any participant experiences any event of abscess, ulceration, or necrosis at the injection site that is considered related to study vaccine administration.
- Two (2) or more participants experience an allergic reaction such as 7eutralizin urticaria (defined as occurring at 3 or more body parts) within 72 hours after study vaccine injection, that is considered related to the vaccine.
- Three (3) or more participants experience a Grade 3 AE (unsolicited and/or clinical laboratory abnormality), in the same Preferred Terms (PT) based on the Medical Dictionary for Regulatory Activities (MedDRA) coding, that lasted at least 48 hours after administration of the vaccine and is considered related to the vaccine. Clinical laboratory abnormalities are not subject to the time window.
- If more than 50% of participants in each cohort in each age group in Phase 1 of the study have a body temp >38.5 Celsius degrees ( C) within 24 hours after the administration of the first injection (as measured by the participants at home and followed up by site via phone), the second injection will not be administered.

**Table S3:** The Immunogenicity laboratory methods

| |
|---|
| **Live-virus Micro-Neutralization Tests (MicroVNT-50)**<br><br>The micro-neutralization assay (MN) is a fundamental test in virology, immunology, vaccine assessment, and epidemiology studies. Since the SARS-CoV-2 or COVID-19 outbreak at the end of December 2019 in China, it has become extremely important to have well-established and validated diagnostic and serological assays for this new emerging virus due to the lack of specific antiviral drugs or vaccines. Since the antibody response of the serum, after a natural SARS-CoV infection remains detectable for a long time, medical authorities in many countries are trying to calculate the percentage of the population that may be protected against the new circulating strain through the assessment of anti-SARS-CoV-2 Immunoglobulin G (IgG) and M (IgM) levels in serum samples. To date, this assay, currently considered the gold-standard is the most specific and sensitive serological assay capable of evaluating and detecting, functional neutralizing antibodies (nAbs).<br><br>The cell-based, Viral NP detection indirect ELISA is detected by using the SARS-CoV/SARS-CoV-2 Nucleocapsid (NP) monoclonal antibody (Sino Biology, USA). Then goat anti-rabbit antibody with horseradish peroxidase (HRP) (Dako, Dennark) conjugate will bind to the primary antibody. When adding peroxidase substrate, the peroxidase (Seracare, USA) will develop deep blue soluble product in dark. The color will be halted by adding 1N HCl solution. After stopping, read by an ELISA reader at wavelength of 450 nm and 620 nm for reacted and unreacted substrate, respectively. The dual O.D.450/620 value is used for calculation of Tissue Culture Infectious Dose (TCID50/ml) titer of the virus. Assay readouts assessed as immune correlates were first expressed in assay values relative to the WHO International Standard for anti-SARS-CoV-2 immunoglobulin The First WHO International Standard for anti-SARS-CoV-2 immunoglobulin is the freeze-dried equivalent of 0.25 mL of pooled plasma obtained from eleven individuals recovered from SARS-CoV-2 infection. The preparation was evaluated in a WHO International Collaborative study. The intended use of the International Standard is for the calibration and harmonization of serological assays detecting anti-SARS CoV-2 neutralizing antibodies. |
| **Pseudovirus neutralization test (psVNT-50)**<br><br>Pseudovirus (PV) neutralization test (psVNT-50), also known as Pseudotyped Virus neutralization assay, is a method used to assess the effect of antibodies (or inhibitors) to block PV-mediated transduction into host cells by directly binding to the SARS-CoV-2 S glycoprotein present on the surface of the PVs. The applications of psVNT-50 as a platform to investigate vaccine efficacy, antigenic properties, and the entry mechanisms of emerging viruses have been utilized in laboratories worldwide. A high correlation between wild-type virus neutralization and psVNT-50 has been demonstrated by several independent studies. The assay was performed on 293T-hACE2 cells expressing human TMPRSS2. Serum samples were serially diluted by2 fold DMEM at a starting dilution of 1:40. Diluted sera were mixed with the same volume of PVs and was incubated for 1 h at 37 °C incubator. Then, the sera-PVs mixtures were added into 96-well plates with 293T-ACE2-TMPRSS2. Plates were incubated at 37°C for 48 h. Then cells were lysed, incubated with luciferase substrate, and the luminescent signal was measured by a microplate reader. The 50% inhibitory concentration (IC50) was calculated using GraphPad Prism. If the fitting value of IC50 is negative (i.e. negative titer), which suggested undetectable neutralization activity, the value was set to baseline. |
| **SARS-CoV-2 RBD and human ACE2 binding inhibition activity or Surrogate viral neutralization test (sVNT).**<br><br>This assay is used for determination of antibodies against SARS-CoV-2 that block the interaction between the receptor binding domain of the viral spike glycoprotein (RBD) with the ACE2 cell surface receptor. The assay detects any antibodies in serum and plasma that inhibit the RBD-ACE2 interaction or Surrogate viral |

| |
|---|
| neutralization test (sVNT) (Genscript, Singapore). First, the samples and controls are pre-incubated with the HRP-RBD to allow the binding of the circulating neutralization antibodies to HRP-RBD. The mixture is then added to the capture plate which is pre-coated with the hACE2 protein. The unbound HRP-RBD as well as any HRP-RBD bound to non-neutralizing antibody will be captured on the plate, while the circulating neutralization antibodies-HRP-RBD complexes remain in the supernatant and get removed during washing. After washing steps, TMB solution is added, making the color blue. By adding Stop Solution, the reaction is quenched, and the color turns yellow. This final solution can be read at 450 nm in a microtiter plate reader. The absorbance of the sample is inversely dependent on the titer of the anti-SARS-CoV-2 neutralizing antibodies. Percent inhibition were calculated by comparing the OD value of sample and negative control. |
| **IgG antibody against S trimer protein of SARS-CoV-2 by enzyme linked immunosorbent assay (ELISA).**<br><br>The Anti-SARS-CoV-2 Antibody IgG Titer Serologic Assay kit (S protein trimer, TAS-K007, Acro Biosystems, USA) employs a standard capture-ELISA format performed according to the manufacturer instruction. , providing a rapid detection of anti-SARS-CoV-2 antibody, IgG in serum by S protein trimer. In brief, the plates were coated with SARS-CoV-2 S protein trimer at 200 ng/well and incubate overnight at 4°C. After excessive washed with 0.5% Tween-20 in TBS (TBST), the plates were blocked with blocking provided in the kit then incubate at 37°C for 1.5 hours. The plates were washed again, serum samples that pre-diluted at 1:50 with dilution buffer were added at 100 µL/well and incubate at 37°C for 1 hour. The unbound antibodies were washed, the 2$^{nd}$ antibody, 100 µL/well of HRP-anti-human IgG (400 ng/mL) were added followed by incubation at 37°C for 1 hour. After washing, 100 µL/well of TMB substrate (BioLegend, USA) was added and incubated for 5 min. The reactions were then stopped with 50 µL of stop solution, 2N sulfuric acid. The absorbance was measured by spectrophotometer at 450 nm and 630 nm using Varioskan microplate reader (ThermoFisher Scientific, Finland). Anti-SARS-CoV-2 IgG level was calculated according to the standard curves of control IgG provided in the kit as well as WHO reference serum (NISB 20/136) |
| **SARS-CoV-2-RBD-specific IgG antibody (SARS-CoV-2 IgG II Quant) Chemiluminescent microparticle immunoassay (CMIA)**<br><br>SARS-CoV-2 anti-RBD IgG antibody binding assays (US Emergency Use Authorization approved Architect, Abbott, Ireland) performed to quantify levels of IgG antibody binding to SARS-CoV-2 RBD of the wild type Wuhan strain and possibly of variants of concern. This includes antibodies with and without neutralising properties. The 1st WHO International Standard for anti-SARS-CoV-2 antibody with known levels of anti-RBD could be used as reference standard (NIBSC, anticipated delivery date December 2020). Low and high titre quality control samples will be used to monitor assay performance. The CMIA assay enables simultaneous quantitative detection of IgG antibodies to three distinct SARS-CoV-2 antigens in human serum. Recently the arbitrary units were bridged to the WHO International Standard and a conversion factor was calculated. Conversion of AU/ml binding antibody readouts to BAU/ml. |

**SARS-CoV-2 Spike Protein IFN-γ T-cell ELISpot vaccine-induced T-cell responses**

The ELISPOT assay is an adaptation of the ELISA, which measures the local concentration of cytokines (i.e., IFN-gamma) that are released from activated CD8+ T cell. In the ELISPOT method, cells that have been stimulated with antigen in vitro are incubated in nitrocellulose-lined micrometer wells, which are pre-coated with anticytokine antibody. After incubation (for several hours or days), the local production of cytokines around "producing cells" can be visualized by adding a second antibody that is labeled with enzyme alkaline phosphatase (ALP) or horseradish peroxidase, and then adding a substrate that will be enzymatically converted into an insoluble colored product. Cytokine-producing cells can then be visualized as "spots".

SARS-CoV-2 Spike Protein IFN-γ T-cell ELISpot vaccine-induced T-cell responses were quantified using an IFN-γ (effector) ELISpot assay established at the Chula VRC. PBMCs were stimulated with SARS-CoV-2 Spike protein peptide pools (Mimotopes, Australia) in plates coated with purified IFN-γ capture antibody. Biotinylated IFN-γ labeled with enzyme ALP detection antibody is added, and signals are amplified by BCIP/NBT substrate solution. Spots were counted on an ELISpot Reader (ImmunoSpot® Analyzer, Germany). Results were expressed as spot forming cells per $10^6$ PBMC. Images of ELISpot plates were kept allowing the spots to be re-counted.

**SARS-CoV-2 spike protein-specific CD4+ and CD8+ T-cell responses and Th1/Th2 polarization responses quantified by intracellular cytokine staining (ICS)**

PBMCs were stimulated with SARS-CoV-2 Spike protein peptide pools (Mimotopes, Australia) in the presence of costimulating antibodies CD28/49d (BD Biosciences) and a cocktail of Golgi Plug and Golgi Stop (BD Biosciences). Following stimulation, cells were washed with staining buffer (PBS with 2% fetal bovine serum [FBS]) and then stained for 20 min at 4°C with the surface markers CD3-PerCP Cy5.5 (OKT3; Biolegend), CD8-AF700 (SK1; Biolegend), and CD4-APC Cy7 (OKT4; Biolegend) in the dark. All cells were fixed for 20 min at 4°C with 2% paraformaldehyde–PBS (Sigma-Aldrich) and subsequently permeabilized. Intracellular staining was done with IL-2- PE (MQ1-17H12; Biolegend), IL-4-PE-DZ594 (MP4-25D2; Biolegend) and IFN-γ–AF647 (4S.B3; Biolegend), for 30 min at room temperature in the dark. After being washed, all cells were resuspended in staining buffer and analyzed on a FACS LSRII flow cytometer with FACS Diva software (Becton Dickinson, San Jose, CA). Data were analyzed using FlowJo 10.4 (Ashland, OR).

**Table S4:** Demographic characteristics of 30 human convalescent serum samples from adults with COVID-19 by severity of COVID-19

| Characteristic | Mild/Moderate<br><br>N=15 | Severe<br><br>N=15 | Overall<br><br>N=30 |
|---|---|---|---|
| **Sex -number (%)** | | | |
| **Female** | 14 (93.3) | 5 (33.3) | 19(63.3) |
| **Age - Mean (SD)** | 29.3(12.4) | 50.4(13.5) | 39.9(16.6) |
| **Min-max** | 14-63 | 21-76 | 14-76 |
| **Ethnicity -number (%)** | | | |
| **Thai** | 15(100%) | 15(100%) | 30(100%) |

Mild/moderate: defined as not required mechanical ventilation

Severe: defined as required mechanical ventilation

**Table S5:** Demographic characteristics of 27 Pfizer/BioNTech mRNA vaccinees

| Characteristic | Overall<br><br>N=27 |
|---|---|
| **Sex -number (%)** | |
| **Female** | 21(77.8) |
| **Age - Mean (SD)** | 34.5(9.4) |
| **Min-max** | 24-57 |
| **Ethnicity -number (%)** | |
| **Malaysian** | 27(100%) |

**Figure S1:** Enrollment and Distribution Adult Cohort

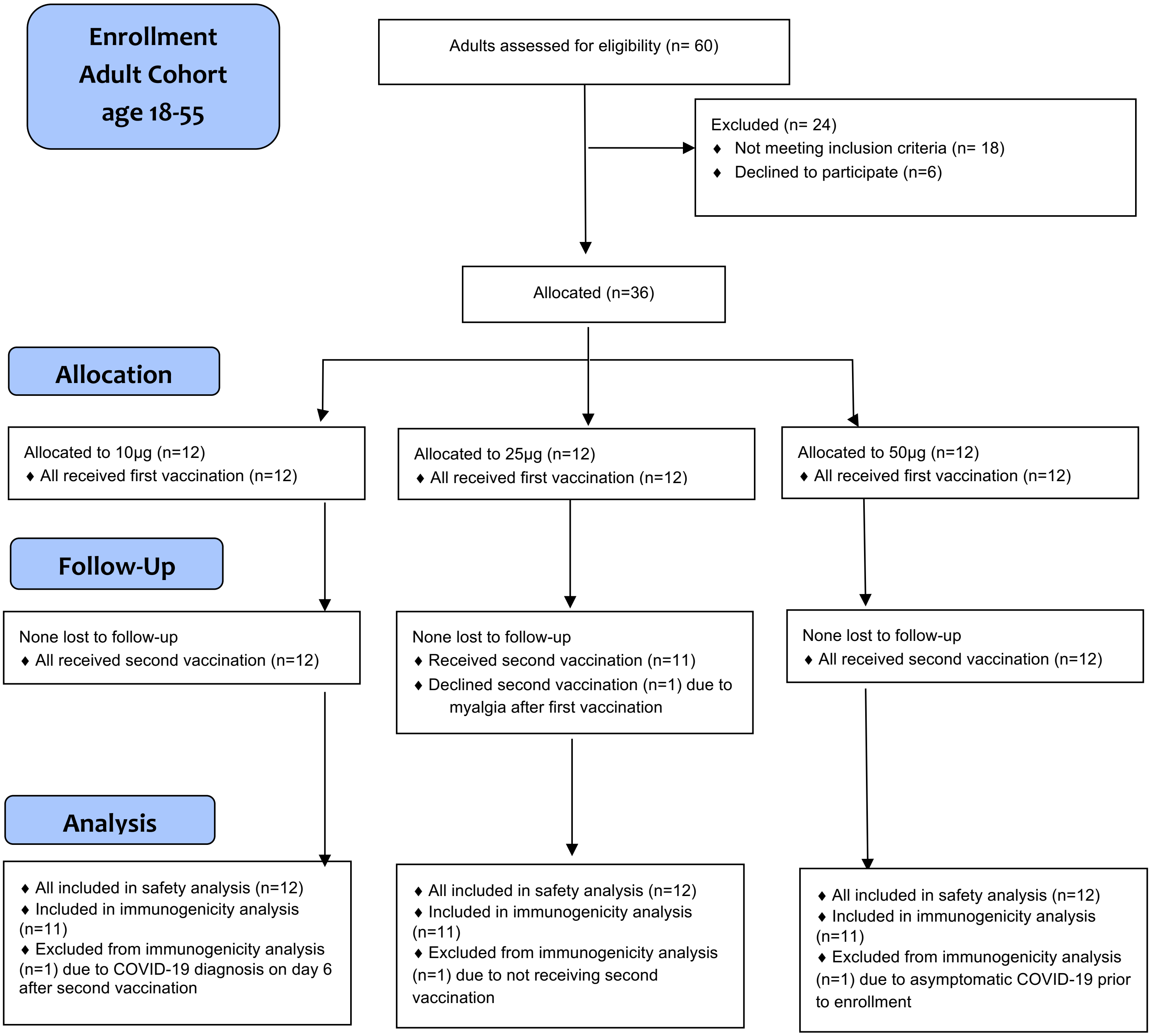

**Figure S2:** Enrollment and Distribution Elderly Cohort

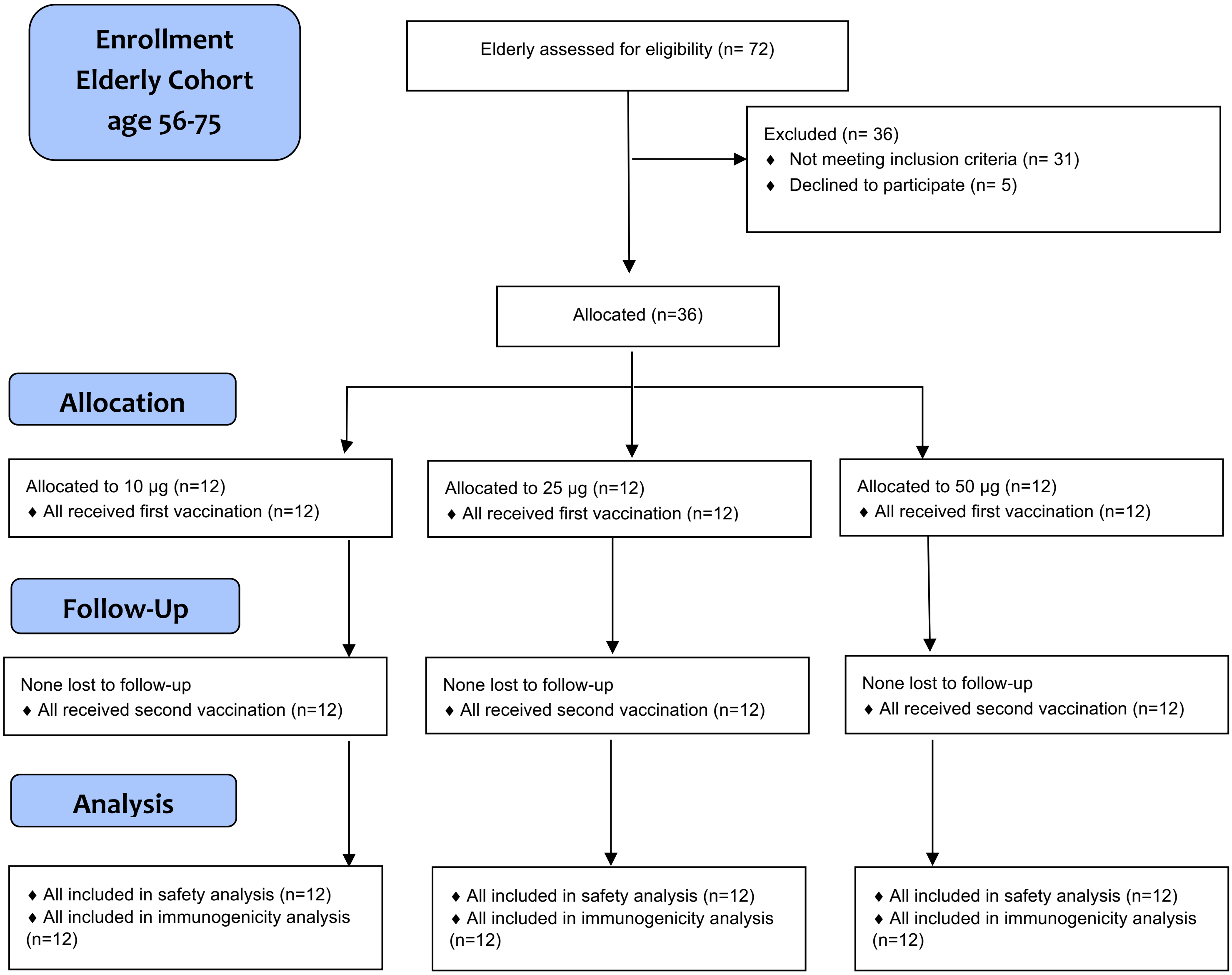

**Table S6**: Comparison of Anti RBD-IgG Antibody (BAU/mL) at Day 29 and 50 Results of ChulaCov19 Vaccine 25 and 50 μg Dose with 10 μg Dose as Reference

| Day of Study | Phase | Dose | N | GMT | | | GMT-Ratio | | | P-value |
|---|---|---|---|---|---|---|---|---|---|---|
| | | | | BAU/mL | 95%CI | | Fold different | 95%CI | | |
| | | | | | LL | UL | | LL | UL | |
| **29** | **Adults** 18-55 yo | 10 | 11 | **135.0** | 49.8 | 366.3 | Ref | Ref | Ref | Ref |
| | | 25 | 11 | **277.1** | 113.5 | 676.5 | **2.05** | 0.67 | 6.28 | 0.20 |
| | | 50 | 11 | **2540.2** | 1308.3 | 4932.1 | **18.81** | 6.15 | 57.56 | **<0.001** |
| | **Elderly** 56-75 yo | 10 | 12 | **107.1** | 50.9 | 225.7 | Ref | Ref | Ref | Ref |
| | | 25 | 12 | **233.6** | 126.7 | 430.9 | **2.18** | 0.91 | 5.23 | 0.08 |
| | | 50 | 12 | **1176.6** | 617.7 | 2241.3 | **10.98** | 4.58 | 26.35 | **<0.001** |
| **50** | Phase 1 :18-55 years | 10 | 11 | **1215.6** | 780.1 | 1894.3 | Ref | Ref | Ref | Ref |
| | | 25 | 11 | **1040.1** | 668.9 | 1617.4 | **0.86** | 0.48 | 1.51 | 0.58 |
| | | 50 | 11 | **2490.1** | 1614.7 | 3840.2 | **2.05** | 1.16 | 3.62 | **0.02** |
| | Phase 1 :56-75 years | 10 | 12 | **845.6** | 502.1 | 1424.2 | Ref | Ref | Ref | Ref |
| | | 25 | 12 | **983.0** | 619.9 | 1558.9 | **1.16** | 0.63 | 2.16 | 0.63 |
| | | 50 | 12 | **2282.0** | 1474.4 | 3532.0 | **2.70** | 1.45 | 5.02 | **0.003** |

GMT: Geometric mean titer, GMTR: Geometric mean titer ratio, 95%CI: 95% confidence interval, LL = lower limit, UL = upper limit, P-value were evaluated by linear regression, ref : reference, Dose 10 μg is reference.

**Figure S3:** Surrogate Viral Neutralizing Antibody assay (sVNT)

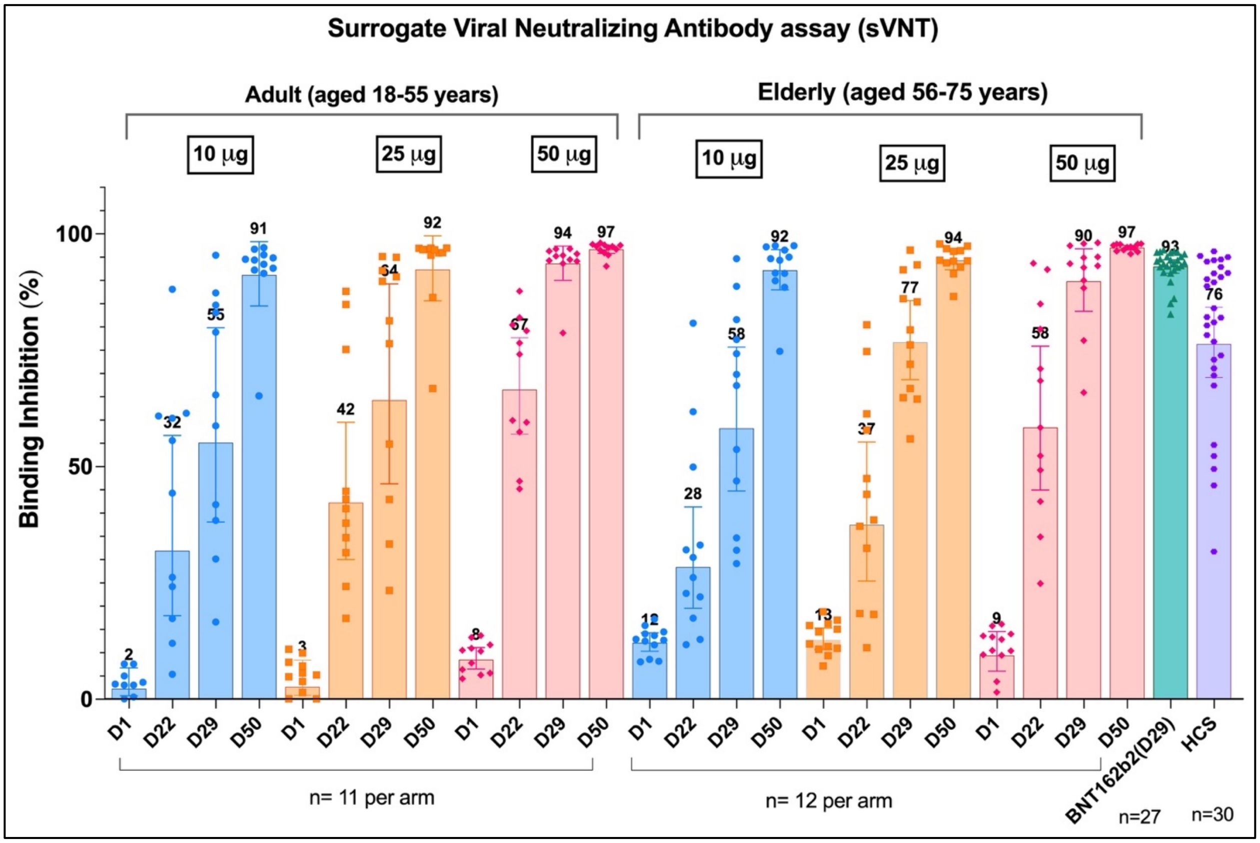

**Table S6**: Comparison of MicroVNT-50 Results of Adult Aged 18-55 against SARS-Cov2 Wild-Type virus and Variants of Concerns between ChulaCov19 vaccine and human Convalescent Serum Panel as a Reference (GMTR= GMT ratio)

| Vaccine, dose | Day 29 of the study | | | Day 50 | | |
|---|---|---|---|---|---|---|
| | **GMT (95%CI)** | **GMFR (95%CI)** | **P-value** | **GMT (95%CI)** | **GMTR (95%CI)** | **P-value** |
| **WT virus (IU/mL): 18-55 years** | | | | | | |
| • 10 µg | 153.6 (59-399.8) | 0.54 (0.23-1.26) | 0.15 | 848.2 (483.3-1488.8) | 2.98 (1.63-5.44) | **0.001** |
| • 25 µg | 195.7 (73.7-519.9) | 0.69 (0.32-1.46) | 0.32 | 736.5 (458.6-1182.7) | 2.59 (1.42-4.72) | **0.003** |
| • 50 µg | 931 (591.9-1464.4) | 3.27 (1.54-6.95) | 0.003 | 1139.9 (853.8-1522) | 4.01 (2.2-7.31) | **<0.001** |
| • Convalescent | **284.6 (195.9- 413.3** | Ref | Ref | **284.6 (195.9- 413.3** | Ref | Ref |
| **WT virus (IU/mL): 55-75 years** | | | | | | |
| • 10 µg | 87 (21.3-169) | 0.31 (0.14-0.68) | 0.005 | 137.9 (86.4-470.1) | 0.48 (0.25-0.92) | 0.03 |
| • 25 µg | 80.5 (22.8-222.8) | 0.28 (0.13-0.61) | 0.002 | 358.4 (511.8-2016.8) | 1.26 (0.67-2.36) | 0.46 |
| • 50 µg | 334.5 (204.2-2005.7) | 1.18 (0.58-2.39) | 0.65 | 594.9 (878.1-5923.6) | 2.09 (1.12-3.91) | 0.02 |
| • Convalescent | 284.6 (195.9-413.3) | Ref | Ref | 284.6 (195.9-413.3) | Ref | Ref |
| **Alpha variant** | | | | | | |
| • 10 µg | **21.3** (9.9-45.7) | 0.09 (0.03-0.24) | **<0.001** | **219.3** (112.1-428.7) | 0.95 (0.45-1.97) | 0.88 |
| • 25 µg | **42.6** (15.9-114.3) | 0.18 (0.07-0.48) | **<0.001** | **411.7** (226.2-749.5) | 1.78 (0.85-3.7) | 0.12 |
| • 50 µg | **264.9** (112-626.5) | 1.14 (0.44-3.01) | 0.78 | **640** (384.3-1065.9) | **2.76** (1.33-5.76) | **0.01** |
| • Convalescent | **231.6** (134.9- 397.5) | Ref | Ref | **231.6** (134.9- 397.5) | Ref | Ref |
| **Beta variant** | | | | | | |
| • 10 µg | **17.6** (9.8-31.6) | 0.04 (0.02-0.09) | **<0.001** | **70.5** (38-130.9) | 0.17 (0.08-0.34) | **<0.001** |
| • 25 µg | **27.4** (14-53.6) | 0.06 (0.03-0.14) | **<0.001** | **109.6** (54.3-221.2) | 0.26 (0.13-0.52) | **<0.001** |
| • 50 µg | **205.9** (86.6-489.2) | 0.49 (0.23-1.02) | 0.06 | **282.1** (142.2-559.6) | 0.67 (0.33-1.34) | 0.25 |
| • Convalescent | **422.2** (318.2-560.3) | Ref | Ref | **422.2** (318.2-560.3) | Ref | Ref |
| **Delta variant** | | | | | | |
| • 10 µg | **18.8** (8.7-40.3) | 0.03 (0.01-0.07) | **<0.001** | **70.5** (31.6-157.2) | 0.11 (0.05-0.25) | **<0.001** |
| • 25 µg | **18.8** (10.2-34.4) | 0.03 (0.01-0.07) | **<0.001** | **150.2** (68.1-331.6) | 0.23 (0.1-0.53) | **<0.001** |
| • 50 µg | **193.3** (77.8-480.2) | 0.3 (0.13-0.71) | **0.001** | **320** (184.4-555.2) | 0.5 (0.22-1.14) | 0.10 |
| • Convalescent | 640 (387.2-1057.9) | Ref | Ref | **640** (387.2-1057.9) | Ref | Ref |
| GMT: Geometric mean titer, GMTR: Geometric mean titer ratio, p-value from linear regression model, ref: reference group | | | | | | |

**Table S7**: Comparison of SARS-COV-2-specific Serum Neutralizing Antibody as Measured By Live-Virus Micro-VNT50- Against Wild-type, Alpha, Beta, and Delta at Day 29 and 50 of ChulaCov19 Vaccine Doses with 10 μg as a Reference.

| Virus | Day | Phase 1 by age | Dose | N | GMT | | | GMFR | | | P-value |
|---|---|---|---|---|---|---|---|---|---|---|---|
| | | | | | MicroVNT50 | 95%CI | | Ratio | 95%CI | | |
| | | | | | | LL | UL | | LL | UL | |
| **WT (IU/mL)** | 29 | 18-55 years | 10 | 11 | **153.6** | 59 | 399.8 | Ref | Ref | Ref | Ref |
| | | | 25 | 11 | **195.7** | 73.7 | 519.9 | 1.27 | 0.43 | 3.77 | 0.65 |
| | | | 50 | 11 | **931** | 591.9 | 1464.4 | 6.06 | 2.05 | 17.91 | 0.002 |
| | | 56-75 years | 10 | 12 | **87** | 39.3 | 192.5 | Ref | Ref | Ref | Ref |
| | | | 25 | 12 | **80.5** | 31.9 | 202.9 | 0.93 | 0.34 | 2.55 | 0.88 |
| | | | 50 | 12 | **334.5** | 185.2 | 604.4 | 3.85 | 1.46 | 10.14 | 0.008 |
| | 50 | 18-55 years | 10 | 11 | **848.2** | 483.3 | 1488.8 | Ref | Ref | Ref | Ref |
| | | | 25 | 11 | **736.5** | 458.6 | 1182.7 | 0.87 | 0.48 | 1.57 | 0.63 |
| | | | 50 | 11 | **1139.9** | 853.8 | 1522 | 1.34 | 0.74 | 2.43 | 0.32 |
| | | 56-75 years | 10 | 12 | **137.9** | 83.2 | 228.5 | Ref | Ref | Ref | Ref |
| | | | 25 | 12 | **358.4** | 224.7 | 571.5 | 2.60 | 1.28 | 5.29 | 0.01 |
| | | | 50 | 12 | **594.9** | 317.6 | 1114.2 | 4.31 | 2.12 | 8.78 | <0.001 |
| **Alpha** | 29 | 18-55 years | 10 | 11 | **21.3** | 9.9 | 45.7 | Ref | Ref | Ref | Ref |
| | | | 25 | 11 | **42.6** | 15.9 | 114.3 | 2.00 | 0.64 | 6.22 | 0.22 |
| | | | 50 | 11 | **264.9** | 112.0 | 626.5 | **12.44** | 4.00 | 38.67 | **<0.001** |
| | | 56-75 years | 10 | 12 | **30.0** | 16.3 | 55.0 | Ref | Ref | Ref | Ref |
| | | | 25 | 12 | **25.2** | 10.6 | 60.0 | 0.84 | 0.27 | 2.67 | 0.76 |
| | | | 50 | 12 | **151.0** | 50.2 | 454.8 | **5.04** | 1.59 | 15.97 | **0.01** |
| **Beta** | 29 | 18-55 years | 10 | 11 | **17.6** | 9.8 | 31.6 | Ref | Ref | Ref | Ref |
| | | | 25 | 11 | **27.4** | 14.0 | 53.6 | 1.55 | 0.61 | 3.93 | 0.34 |
| | | | 50 | 11 | **205.9** | 86.6 | 489.2 | 11.68 | 4.62 | 29.53 | **<0.001** |
| | | 56-75 years | 10 | 12 | **10.6** | 9.3 | 12.0 | Ref | Ref | Ref | Ref |
| | | | 25 | 12 | **15.9** | 9.9 | 25.5 | 1.50 | 0.72 | 3.10 | 0.27 |
| | | | 50 | 12 | **37.8** | 16.5 | 86.4 | 3.56 | 1.72 | 7.37 | **0.001** |
| **Delta** | 29 | 18-55 years | 10 | 11 | **18.8** | 8.7 | 40.3 | Ref | Ref | Ref | Ref |
| | | | 25 | 11 | **18.8** | 10.2 | 34.4 | 1.00 | 0.37 | 2.71 | 0.99 |
| | | | 50 | 11 | **193.3** | 77.8 | 480.2 | 10.29 | 3.79 | 27.92 | <0.001 |
| | | 56-75 years | 10 | 12 | **10.0** | 10.0 | 10.0 | Ref | Ref | Ref | Ref |
| | | | 25 | 12 | **15.0** | 8.7 | 25.9 | 1.50 | 0.73 | 3.06 | 0.26 |
| | | | 50 | 12 | **44.9** | 20.8 | 97.0 | 4.49 | 2.20 | 9.16 | **<0.001** |

GMT: Geometric mean titer, GMTR: Geometric mean titer rise ratio, 95%CI: 95% confidence interval, LL = lower limit, UL = upper limit, P-value were evaluated by linear regression, ref; reference, Dose 10 μg is reference.

**Table S8:** Comparison of Pseudovirus Neutralizing Antibody (psVNT-50) Results of ChulaCov19 Vaccine in Adult Cohort with Pfizer /BNT Vaccine as a Reference (ChulaCov19 n=11/dose group, Pfizer/BNT panel, n=27)

| Vaccine, dose | Day 29 | | P-value |
|---|---|---|---|
| | GMT (95%CI) | GMTR (95%CI) | |
| **WT virus** | | | |
| • 10 µg | **27.5** (2.5-297.9) | **0.07** (0.02-0.28) | **<0.01** |
| • 25 µg | **107.5** (20.8-556.2) | 0.28 (0.07-1.11) | 0.07 |
| • 50 µg | **1284.5** (662.6-2490.1) | **3.3** (0.83-13.22) | 0.09 |
| • Pfizer/BioNTech | **388.6** (289.4-521.9) | Ref | Ref |
| **Alpha variant** | | | |
| • 10 µg | **21.8** (3.3-144) | **0.1** (0.03-0.3) | **<0.01** |
| • 25 µg | **156.7** (51.6-475.9) | 0.73 (0.23-2.31) | 0.58 |
| • 50 µg | **964.4** (442.9-2100) | **4.47** (1.53-13.08) | **0.01** |
| • Pfizer/BioNTech | **215.6** (165.3-281.1) | Ref | Ref |
| **Beta variant** | | | |
| • 10 µg | **2.2** (0.4-11.1) | **0.08** (0.01-0.42) | **<0.01** |
| • 25 µg | **1.1** (0-77.2) | **0.04** (0-0.36) | **0.01** |
| • 50 µg | **343.4** (195.8-602.4) | **11.94** (2.24-63.68) | **0.01** |
| • Pfizer/BioNTech | **28.7** (9.3-88.7) | Ref | Ref |
| **Gamma variant** | | | |
| • 10 µg | **2.5** (0.1-47.2) | **0.03** (0.01-0.1) | **<0.01** |
| • 25 µg | **58.4** (15.1-225.1) | 0.66 (0.16-2.69) | 0.56 |
| • 50 µg | **371.8** (216.6-638.1) | **4.23** (1.49-12.01) | **0.01** |
| • Pfizer/BioNTech | **87.8** (66-116.8) | Ref | Ref |
| **Delta variant** | | | |
| • 10 µg | **11.8** (0.7-199.6) | **0.05** (0.01-0.19) | **<0.01** |
| • 25 µg | **134.5** (48.9-370.3) | 0.52 (0.13-2.11) | 0.35 |
| • 50 µg | **976.8** (400.5-2382.3) | **3.75** (0.92-15.35) | 0.07 |
| • Pfizer/BioNTech | **260.2** (168.9-400.5) | Ref | Ref |
| GMT: Geometric mean titer, GMTR: Geometric mean ratio, p-value from linear regression model, ref: reference group | | | |

**Figure S4** Pseudovirus neutralizing antibody results against Omicron variant of Adults and Elderly vaccinated with ChulaCov19 vaccine at 50 μg dose

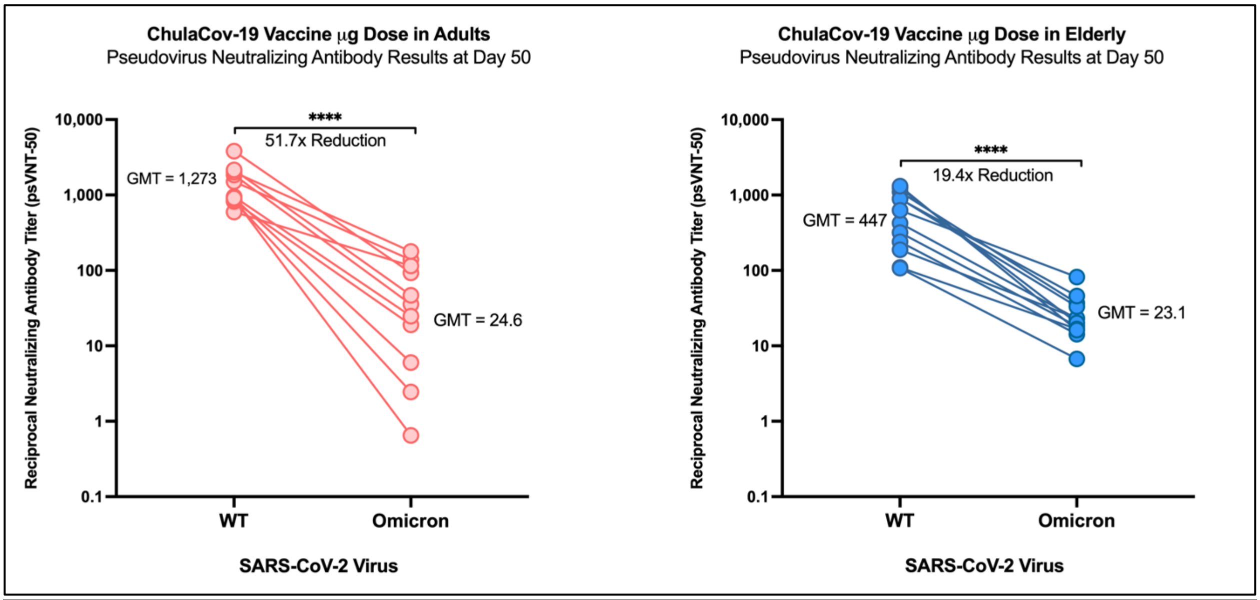

**Table S9:** Comparison of SARS-Cov2-Spike-specific-IFN-γ-ELISPOT T Cell responses at Day 29 of ChulaCov19 Vaccine Doses with 10 μg as a Reference.

| Phase 1 Age group | ChulaCov19 Dose μg | N | GM | | | GMR | | | P-value |
|---|---|---|---|---|---|---|---|---|---|
| | | | ELISpot SFC/1 million PBMCs | 95%CI | | Ratio | 95%CI | | |
| | | | | LL | UL | | LL | UL | |
| 18-55 years | 10 | 11 | **1199.9** | 760.1 | 1894.2 | Ref | Ref | Ref | Ref |
| | 25 | 11 | **2354.0** | 1658.1 | 3341.9 | **1.96** | 1.19 | 3.24 | **0.01** |
| | 50 | 11 | **1875.2** | 1332.5 | 2638.9 | 1.56 | 0.95 | 2.58 | 0.08 |
| 56-75 years | 10 | 12 | **635.3** | 369.6 | 1091.2 | Ref | Ref | Ref | Ref |
| | 25 | 12 | **870.1** | 469.0 | 1614.1 | 1.37 | 0.71 | 2.64 | 0.34 |
| | 50 | 12 | **1719.5** | 1286.2 | 2298.8 | **2.71** | 1.40 | 5.23 | **0.004** |

GM: Geometric mean, GMR: Geometric mean ratio, 95%CI: 95% confidence interval, LL = lower limit, UL = upper limit, P-value were evaluated by linear regression, ref: reference, Dose 10 μg is reference.

**Table S10**: Comparison of percent of SARS-Cov2-Spike-specific-CD4+ T-cell responses and Th1/Th2 ratio measured at day 29 by intracellular cytokine-staining assays

| T-cell | Phase1 | Dose µg | N | GM | | | GM Ratio | | | P-value |
|---|---|---|---|---|---|---|---|---|---|---|
| | | | | % Positive | 95%CI | | Ratio | 95%CI | | |
| | | | | | LL | UL | | LL | UL | |
| **IFNγ** | **Adults** 18-55 yo | 10 | 11 | **0.09** | 0.04 | 0.19 | Ref | Ref | Ref | Ref |
| | | 25 | 11 | **0.26** | 0.15 | 0.47 | **3.08** | 1.43 | 6.63 | **0.01** |
| | | 50 | 11 | **0.22** | 0.17 | 0.30 | **2.63** | 1.23 | 5.66 | **0.02** |
| | **Elderly** 56-75 yo | 10 | 12 | **0.06** | 0.03 | 0.13 | Ref | Ref | Ref | Ref |
| | | 25 | 12 | **0.12** | 0.07 | 0.20 | **2.13** | 0.95 | 4.79 | 0.07 |
| | | 50 | 12 | **0.16** | 0.10 | 0.26 | **2.79** | 1.24 | 6.26 | 0.02 |
| **IL-2** | **Adults** 18-55 yo | 10 | 11 | **0.13** | 0.09 | 0.19 | Ref | Ref | Ref | Ref |
| | | 25 | 11 | **0.32** | 0.24 | 0.44 | **2.50** | 1.67 | 3.74 | **<0.001** |
| | | 50 | 11 | **0.31** | 0.26 | 0.38 | **2.43** | 1.63 | 3.63 | **<0.001** |
| | **Elderly** 56-75 yo | 10 | 12 | **0.13** | 0.10 | 0.18 | Ref | Ref | Ref | Ref |
| | | 25 | 12 | **0.19** | 0.14 | 0.26 | **1.42** | 0.97 | 2.08 | 0.07 |
| | | 50 | 12 | **0.21** | 0.16 | 0.29 | **1.60** | 1.09 | 2.35 | **0.02** |
| **Th1/Th2** | | **Dose** µg | | **GM Th1/Th2** | **95% CI** | | | | | |
| | | | | | LL | UL | | | | |
| **IFNγ/IL-4** | **Adults** 18-55 yo | 10 | 11 | **1.85** | 1.00 | 3.39 | Ref | Ref | Ref | Ref |
| | | 25 | 11 | **3.08** | 1.71 | 5.54 | **1.67** | 0.84 | 3.30 | 0.14 |
| | | 50 | 11 | **3.28** | 2.32 | 4.64 | **1.78** | 0.88 | 3.58 | 0.10 |
| | **Elderly** 56-75 yo | 10 | 12 | **1.04** | 0.50 | 2.16 | Ref | Ref | Ref | Ref |
| | | 25 | 12 | **1.19** | 0.70 | 2.02 | **1.15** | 0.52 | 2.55 | 0.73 |
| | | 50 | 12 | **2.51** | 1.45 | 4.34 | **2.42** | 1.09 | 5.39 | **0.03** |
| **IL-2/IL-4** | **Adults** 18-55 yo | 10 | 11 | **2.90** | 2.13 | 3.96 | Ref | Ref | Ref | Ref |
| | | 25 | 11 | **3.74** | 2.55 | 5.50 | **1.29** | 0.78 | 2.13 | 0.31 |
| | | 50 | 11 | **4.42** | 2.77 | 7.05 | **1.52** | 0.91 | 2.55 | 0.10 |
| | **Elderly** 56-75 yo | 10 | 12 | **2.42** | 1.80 | 3.27 | Ref | Ref | Ref | Ref |
| | | 25 | 12 | **1.86** | 1.37 | 2.53 | **0.77** | 0.49 | 1.21 | 0.25 |
| | | 50 | 12 | **3.39** | 2.20 | 5.21 | **1.40** | 0.88 | 2.21 | 0.15 |

GM: Geometric mean, GMR: Geometric mean ratio, 95% confidence interval, LL = lower limit, UL = upper limit p-value from linear regression model
